# Adaptive time-dependent priors and Bayesian inference to evaluate SARS-CoV-2 public health measures validated on 31 countries

**DOI:** 10.1101/2020.06.10.20126870

**Authors:** Hugues Turbé, Mina Bjelogrlic, Arnaud Robert, Christophe Gaudet-Blavignac, Jean-Philippe Goldman, Christian Lovis

**Author notes:** **Correspondence:** Hugues Turbé.

## Abstract

With the rapid spread of the SARS-CoV-2 virus since Fall 2019, public health confinement measures to contain the propagation of the pandemic are taken. Our method to estimate the reproductive number using Bayesian inference with time-dependent priors enhances previous approaches by considering a dynamic prior continuously updated as restrictive measures and comportments within the society evolve. In addition, to allow direct comparison between reproductive number and introduction of public health measures in a specific country, the infection dates are inferred from daily confirmed cases and death with the mean time between a case being declared as positive and its death estimated on 1430 cases at 10.7 days. The evolution of the reproductive rate in combination with the stringency index is analyzed on 31 European countries. We show that most countries required tough state interventions with a stringency index equal to 83.6 out of 100 to reduce the reproductive number below one and control the progression of the epidemic. In addition, we show a direct correlation between the time taken to introduce restrictive measures and the time required to contain the spread of the epidemic with a median time of 8 days. Our analysis reinforces the importance of having a fast response with a coherent and comprehensive set of confinement measures to control the epidemic. Only combinations of non-pharmaceutical interventions (NPIs) have shown to be effective.

## 1 Introduction

Since being first observed in Wuhan in late 2019, the outbreak of the 2019 SARS-CoV-2 virus is strongly affecting societies and economies. The transmission rate, pressure on the healthcare system and lack of effective treatment lead countries to take public health measures to limit the spread of the virus. The confinement measures range from banning gatherings to complete lockdowns and closing borders ^1,2^. Additional measures include individual protection with various levels of mask wearing injunctions, and contact tracing with quarantine. This work has focused on developing reliable modelling approaches to evaluate the impact of public health measures. Our method is based on analyzing the reporting of European countries to evaluate the temporal influence of non-pharmaceutical interventions (NPIs) on the effective reproduction number R_t_. The effective reproduction number aims to quantify the number of secondary infections caused by an individual over the time at which this person is infected. It is important to make the distinction between the effective and basic reproduction rate. The basic reproduction R_0_ refers to the evolution of the disease when the population is fully susceptible to the disease while the effective reproductive rate R_t_ factors numerous parameters, such as the susceptible population, the transmission, the immunity acquired within the population, amongst others ^3^. The reproduction rate is a key parameter to evaluate the evolution of an epidemic. Any value below one indicates that the spread is decreasing, any value above one indicates that the spread is increasing. The reproductive number allows a direct comparison of the epidemiologic profiles observed in different cohorts of population, such as specific risk factors driven cohorts, countries, etc. with distinct characteristics (population, testing methods, etc). It allows thus to consider temporality and populational or cohorts characteristics.

Numerous methods have been developed to compute the reproductive rate and its evolution over time ^4^ with the aim of identifying the most influential parameters and predicting the development of an epidemic in a given environment. Initial methods derived the reproductive rate from transmission model similar to the SIR model ^5–9^. However, these models require strong assumptions about the epidemiology of the disease. Later models, including the Wallinga and Teunis approach^10^, use a likelihood-based estimation procedure to reconstruct infection patterns. These methods have shown large variations when using daily data^11^. Most approaches aiming at correcting these fluctuations appeared to be sensitive to smoothing parameters^11,12^. An additional method to mitigate these drawbacks that is very robust to underreporting was later developed by Cori et al.^13^

Since the start of the COVID-19 pandemic, various studies have looked at the impact of public health interventions on the evolution at regional or national level. The first studies, on data from China, proving the impact of NPI strategies to reduce the reproductive rate used mechanistic transmission models to obtain the reproductive number, with the drawbacks associated with these models^14,15^. Further studies focused on how state interventions prevented ICU capacity to be overwhelmed as well as their impact on fatalities in the UK^16^, Germany^17^ and France^18^. While these researches focused on individual country, a recent study aimed to demonstrate the impact of non-pharmaceutical interventions in 11 European countries^19^. This study assumed that the various interventions had the same effect across countries and that their impact was independent of the timing of the measure. In addition, this study assumed the reproductive number to be fixed between the different measures. However, a recent research shows that community changes also play a role in slowing the evolution of the virus^20^.

The aim of this work is to extend previous researches by focusing on the effects of state interventions in 31 European countries. As the evolution of the reproductive number is a function of at least three important parameters: the type of the restrictive measures; the effect of these measures and changes in behaviors with specific societal properties and the size of various compartmental cohorts involved, we do not aim to quantify the effect of each measure. Instead, we aim to show how these combined interventions and their temporality have influenced the spread of the virus.

## 2 Material and methods

The impact of public health measures to control SARS-CoV-2 is evaluated by analyzing the reproductive number computed on incidence data. Our methodology is based on estimates of incubation time, onset to confirmed, and onset to death distributions. The estimation of the reproductive rate is a variation of the one proposed by Cori et al.^13^. While their method assumes a constant gamma distribution for the prior distribution, the presented model takes advantage of the information gained in time by updating the prior distribution for each window with the previous posterior.

### 2.1 Determining incubation time, onset to confirmed, and onset to death distributions

The method described below allows to compute the reproductive number without developing a transmission model and hence only requires a hypothesis on the serial interval. The serial interval measures the period between the symptoms onset in a primary care and the symptoms onset in a secondary case infected by the primary case. The number of new infections on a given day can be estimated as follows:

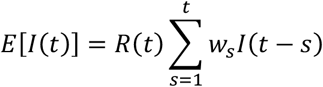

where *w*_*s*_ is the serial interval. The distribution of *w*_*s*_ for the SARS-CoV-2 virus was found to have a mean of 4.8 days and a standard deviation of 2.3 days ^21^.

Given the time at which an infection *I*(*t*) occurred is not available, the number of confirmed cases and deaths on a given day are used as proxies, keeping in mind that the developed method is able to deal with underreporting. A gamma distribution with a median incubation period at 4.4 days from confirmed infection and diagnosis outside of the epicenter of Hubei Province, China, based on official reports from governmental institutes was derived ^22^. The mean and deviation were then obtained by fitting a gamma distribution to the quantile provided in this study. The period between the onset of the symptoms and a case being confirmed in Switzerland was estimated to 5.6 days^23^. Incubation, onset to confirmed, and onset to death periods were assumed to be gamma distributed and their mean and standard deviation are summarized in table 1, where the onset refers to the symptoms onset.

**Table 1:** Incubation and waiting time distribution

| Period | Mean [days] | Standard deviation [days] |
| --- | --- | --- |
| Incubation <sup>22)</sup> | 4.6 | 2.6 |
| Onset to confirmed <sup>23</sup> | 5.6 | 4.2 |
| Onset to death (our study) | 15.3 | 6.7 |

From the latter periods it is possible to calculate a posterior distribution of R_t_ based on the inferred infection dates extracted from the confirmed cases and deaths reported. For the daily cases declared (incidence), a shift following a gamma distribution between the defined cases (confirmed or dead) and the time of infection is randomly generated. For each case, the new date of infection is generated by subtracting the shift to the reported date. This procedure is performed iteratively with the mean of daily simulated number of infections stored. This approach increases the robustness of our method by naturally smoothing the data. This step is critical in order to deal with data unregularly reported.

### 2.2 Correcting the number of infections

In addition, the infections for the most recent days are corrected (Scire et al.^23^) to factor delayed reporting:

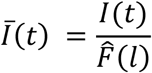

where 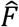 is the cumulative distributive function of the period between an infection and a case being reported as positive or dead, *l* is the time between t and the last reported case, *Ī*(*t*) and *I*(*t*) are respectively the corrected and initial infections which took place on a given day.

### 2.3 Estimation of the reproductive number using Bayesian inference with time-dependent priors

The method presented in this report is a variation of the one proposed by Cori et al.^13^. Assuming the incidence at time t *I*_*t*_ is Poisson distributed so that the likelihood of the incidence *I*_*t*_ given *R*_*t*_ and conditional on previous incidences *I*_0_, …, *I*_*t*−1_:

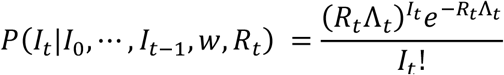

with 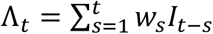 where *w*_*s*_ is the serial interval.

The posterior of the reproductive number *R*_*t*_ conditional on previous incidences is:

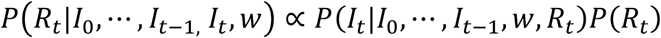

While the method developed by Cori et al.^13^ assumes a constant gamma distribution for the prior distribution, the presented model takes advantage of the information gained in time by updating the prior distribution for each window with the previous posterior:

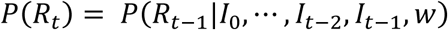

The 95% CI is then derived by computing the 2.5% and 97.5% quantiles.

The reproductive number R_t_ based on the confirmed cases is reported up to 9 days before the last date at which results are available. This corresponds to the median time for confirmed cases to be reported. Using the same method, R_t_ based on the cases reported as dead is reported up to 19 days before the last day on which deaths were reported for a given country.

### 2.4 Inclusion and exclusion criteria

The above described methodology is evaluated on European countries, with a full dataset available for 33 European countries. For our analysis, Russia and Ukraine were removed from our dataset as the reported daily death are still increasing for these two countries when we are interested in countries which have successfully contained the evolution of the pandemic before the 23rd of May 2020. We were therefore left with a set of 31 European countries.

### 2.5 Data Sources and Availability

- R_t_ was estimated using incidence data for confirmed cases and death published in the *COVID-19 Data Repository* ^24^.
- The daily incidence for the death cases per million people was retrieved from Our World in Data ^25^.
- Data related to the period between a positive test and the death of an individual were retrieved from: Swiss Federal Office of Public Health (FOPH)^26^.
- Data regarding the various state interventions were retrieved from the *Coronavirus government response tracker* (OxCGRT) developed by the Blavatnik School of Government (Hale et al. 2020). The stringency index provided in this dataset tracks government’s policies and interventions across different categories and provides a score between 0 and 100 evaluating the overall stringency of the measures taken in a given country ^28^.

## 3 Results

### 3.1 Determining the period between a positive case and death

In this study, the period between a case being reported positive and the death of the individual was extracted from 1430 cases provided by the Swiss Federal Office of Public Health (FOPH). Our result provides a distribution on a much larger dataset than the one built which used between 24 and 33 cases ^22,29^. Three different distributions were tested: lognormal, Weibull and gamma with the Akaike Information Criterion (AIC) being used to identify the best distribution. The results are summarized in Table 3.

**Table 2:**
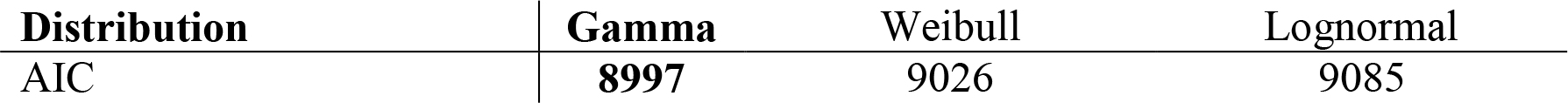
Distribution model and AIC for test to death period

**Table 3:**
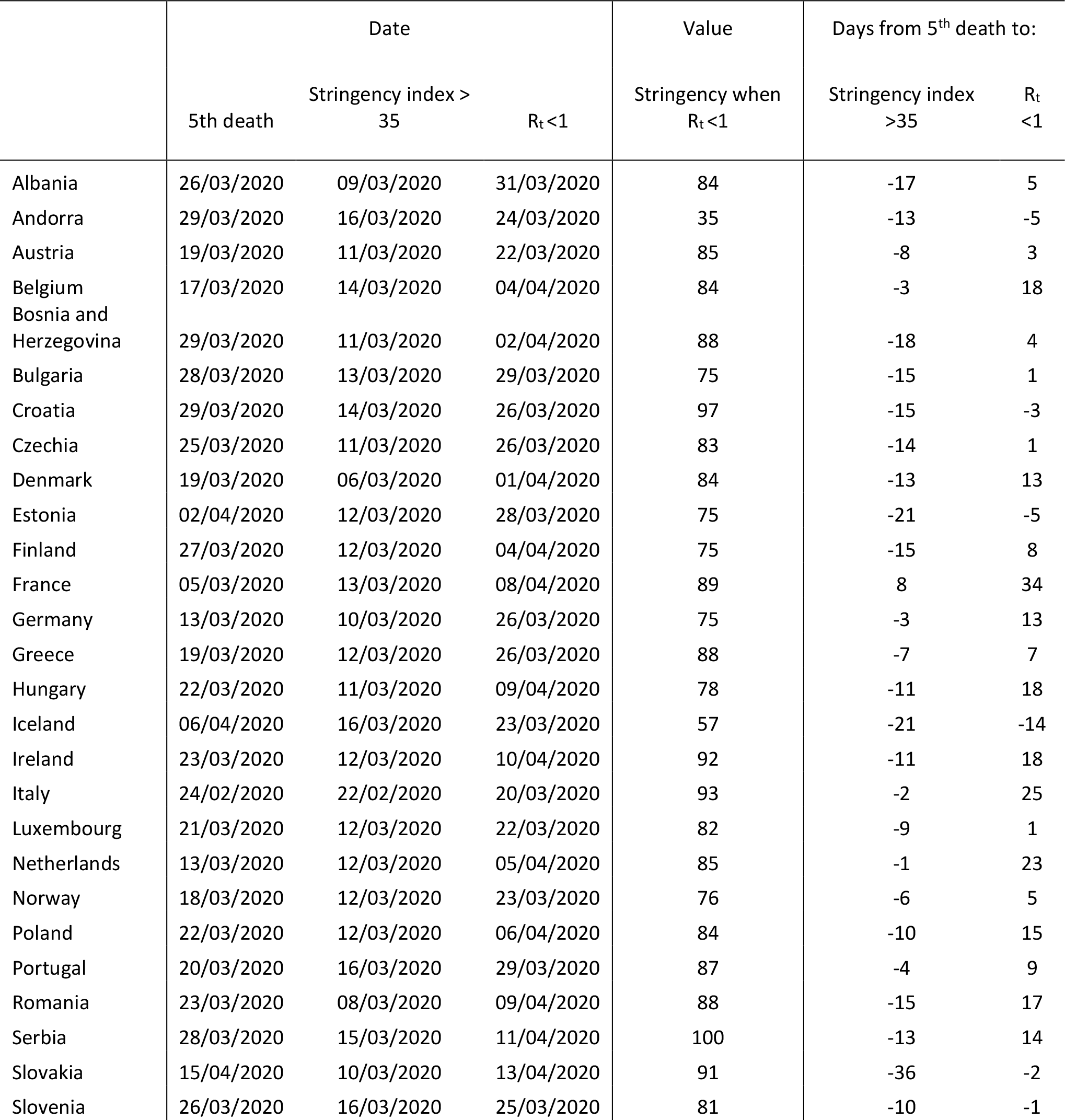
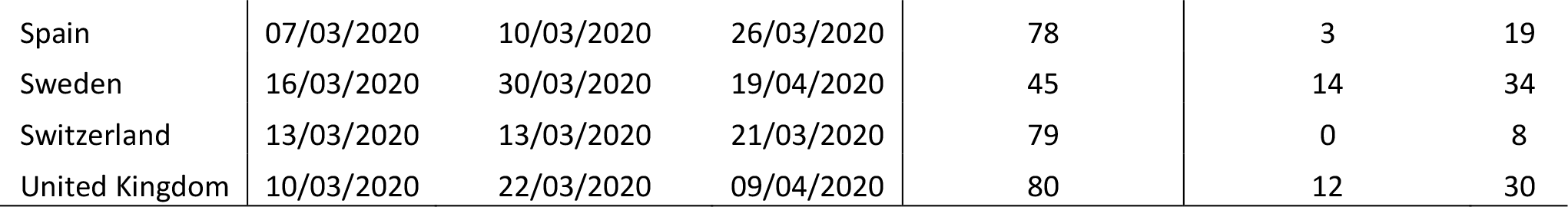
List of countries along dates characterizing the evolution of the epidemic

A gamma distribution with a mean and a standard deviation equal respectively to 10.7 and 6.73 days was found to best fit the data from infection to death. The distribution along the extracted data are shown in Figure 1. This distribution was then combined with the incubation period derived by Linton et al.^22^ to obtain the period between onset and death shown in Table 1.

**Figure 1:**
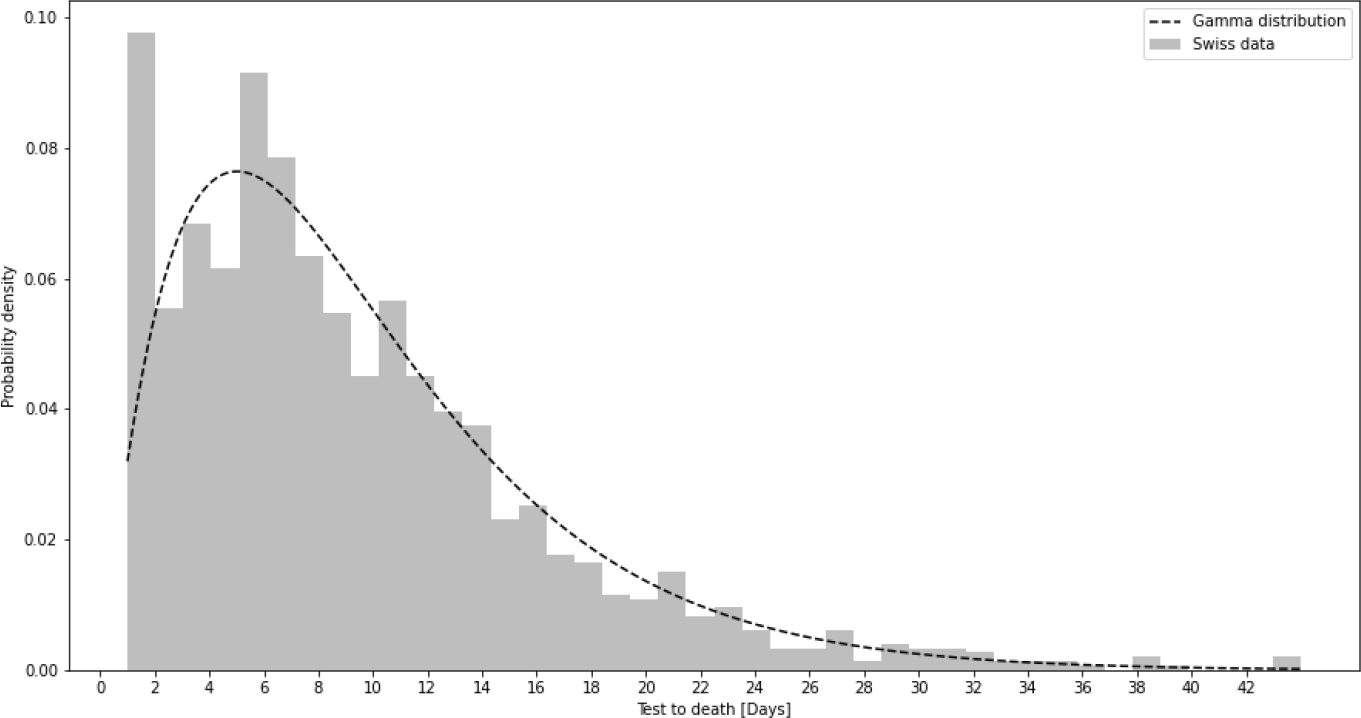
Distribution of the period between a positive test and the death of the patient

### 3.2 Evaluating the reproductive number from incidence data of 31 countries

The evolution of the reproductive rate in Austria is shown in Figure 2. The daily observed confirmed cases and deaths are shown in the top part with blue and violet bars respectively. The same color code is applied in the top part for the estimated daily infections date inferred from these cases and in the middle plot for the derived reproductive rates. These results are shown along the evolution of the stringency index displayed in the bottom plot. Austria is a good analysis case. Indeed we can observe that the reproductive rate started to decline before the introduction of restrictive measures between March 13^th^ and 17^th^, but the reduction was intensified by these measures. The reproductive rate then plateaued at around 0.65 during the lockdown. Recently, the reproductive rate has been oscillating around one. This last phase shows the difficulty to reduce the number of cases below a certain threshold with the emergence of localized clusters, whose identification will be critical to avoid a second wave.

**Figure 2:**
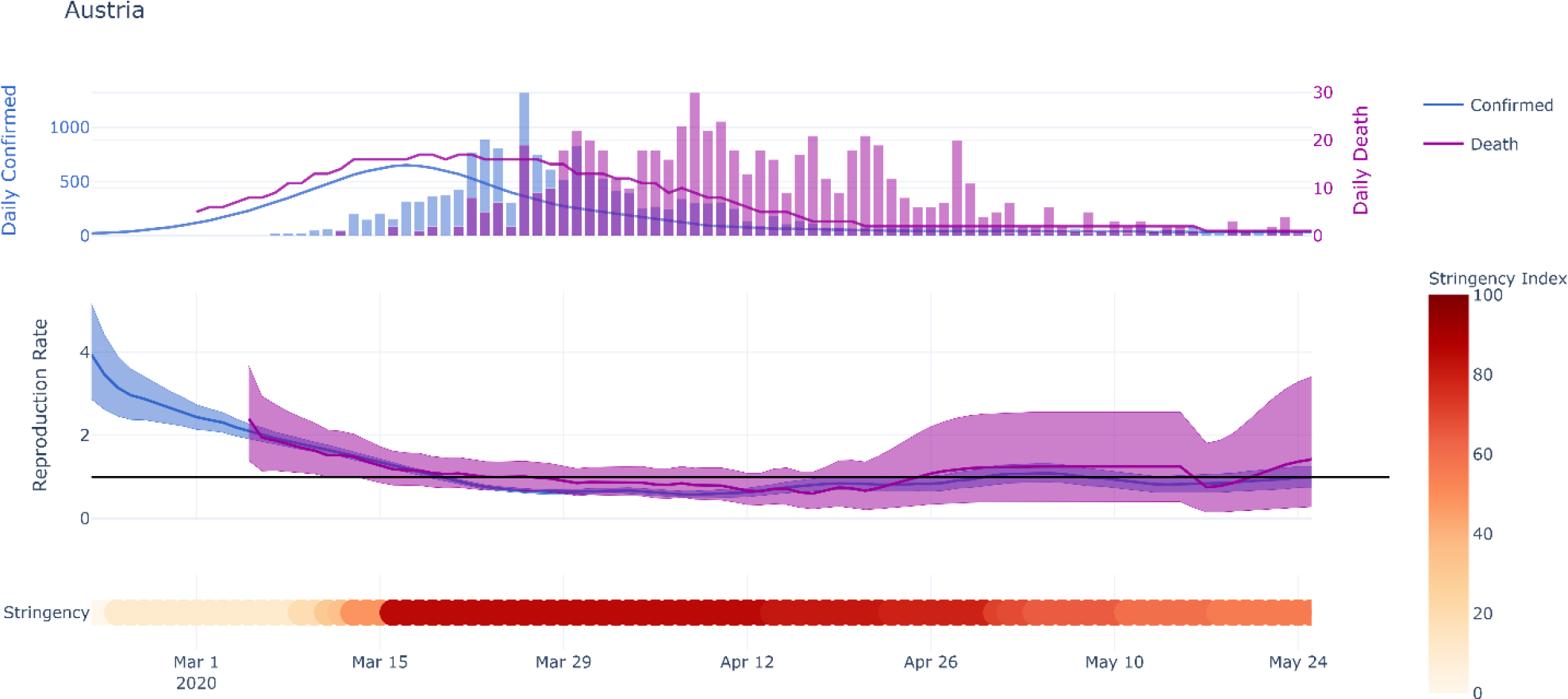
Evolution of the reproductive rate (mean and 95% confidence interval) in Austria. Top: reported deaths and confirmed cases shown with bars, inferred estimated infection distributions shown as solid lines. Middle: the solid lines represent the mean of the estimated reproductive rate and the shaded area the 95% CI (see Methods). Bottom: the evolution of the stringency index is plotted.

The stringency index developed as part of the OxCGRT project^28^ was used to assess the role of state interventions in controlling the epidemic. This index was compared with the evolution of the reproductive rate, rather than the incidence of confirmed or dead cases. It helps comparing countries that have heterogeneous testing or reporting policies. While the reproductive rate is also subject to variations in these policies, it depends on the change in confirmed and death cases, therefore allowing comparison between countries with different policies. For each country, the public health measures and the stringency index are analyzed when reproductive number based on the confirmed cases dropped below one. The hypothesis is that it can help identifying the most efficient set of public health measures. The stringency indexes of the analyzed countries are presented in Figure 3. When countries managed to reduce their reproductive rate below one, they had a mean stringency index of 83.6 out of 100 with a standard deviation of 13.5. When R_t_ dropped below one, the median severity of the measures along their individual severity out of 100 for each category defined in the OxCGRT dataset was the following: a) School closed (100/100); b) Non-essential economic activities closed (100/100); c) Public events were cancelled (100/100); d) Gathering of more than 10 people banned (100/100); e) Mandatory at home policy with minimal exceptions (67/100); f) Movements in the country were restricted (100/100).

**Figure 3:**
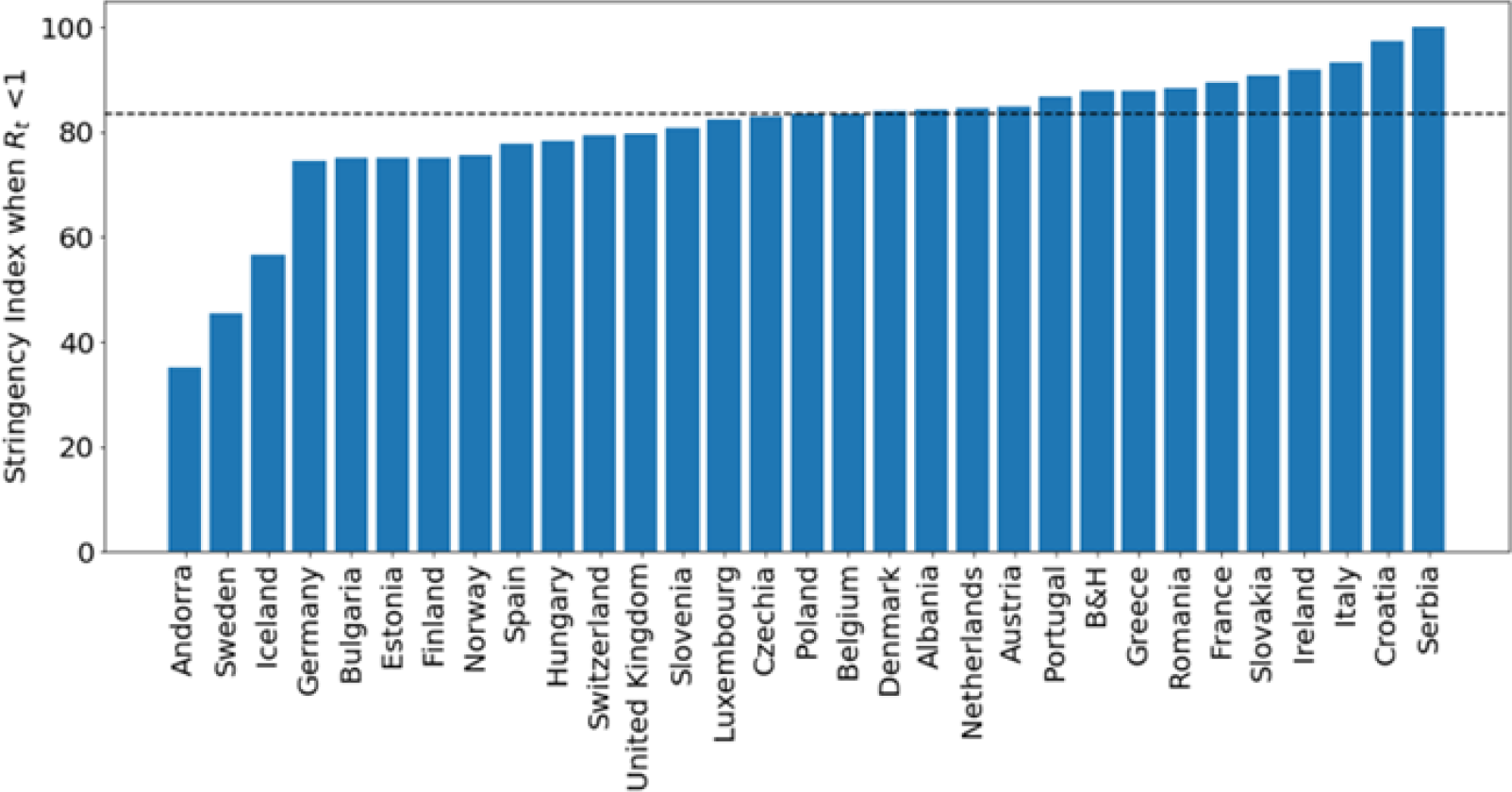
Stringency index per country when Rt dropped below 1. The median value is indicated with the dotted line.

In order to assess the impact of taking restrictive measures early in the crisis, the time taken to introduce initial restrictive measures was compared to the period taken to control the epidemic. The time until the introduction of restrictive measure was defined as the period between the fifth death in a given country and the stringency index reaching a score of 35. The stringency index threshold at 35 corresponds to the lowest score observed when a country reached a R_t_ smaller than one which was observed for Andorra. The time required to control the epidemic was then defined as the period between the fifth death and the reproductive rate based on the confirmed cases dropping below one. The results along a linear regression are presented in Figure 4. A Pearson correlation coefficient of 0.762 was found between the two variables indicating that there is indeed a positive benefit of taking early measures to control the epidemic.

**Figure 4:**
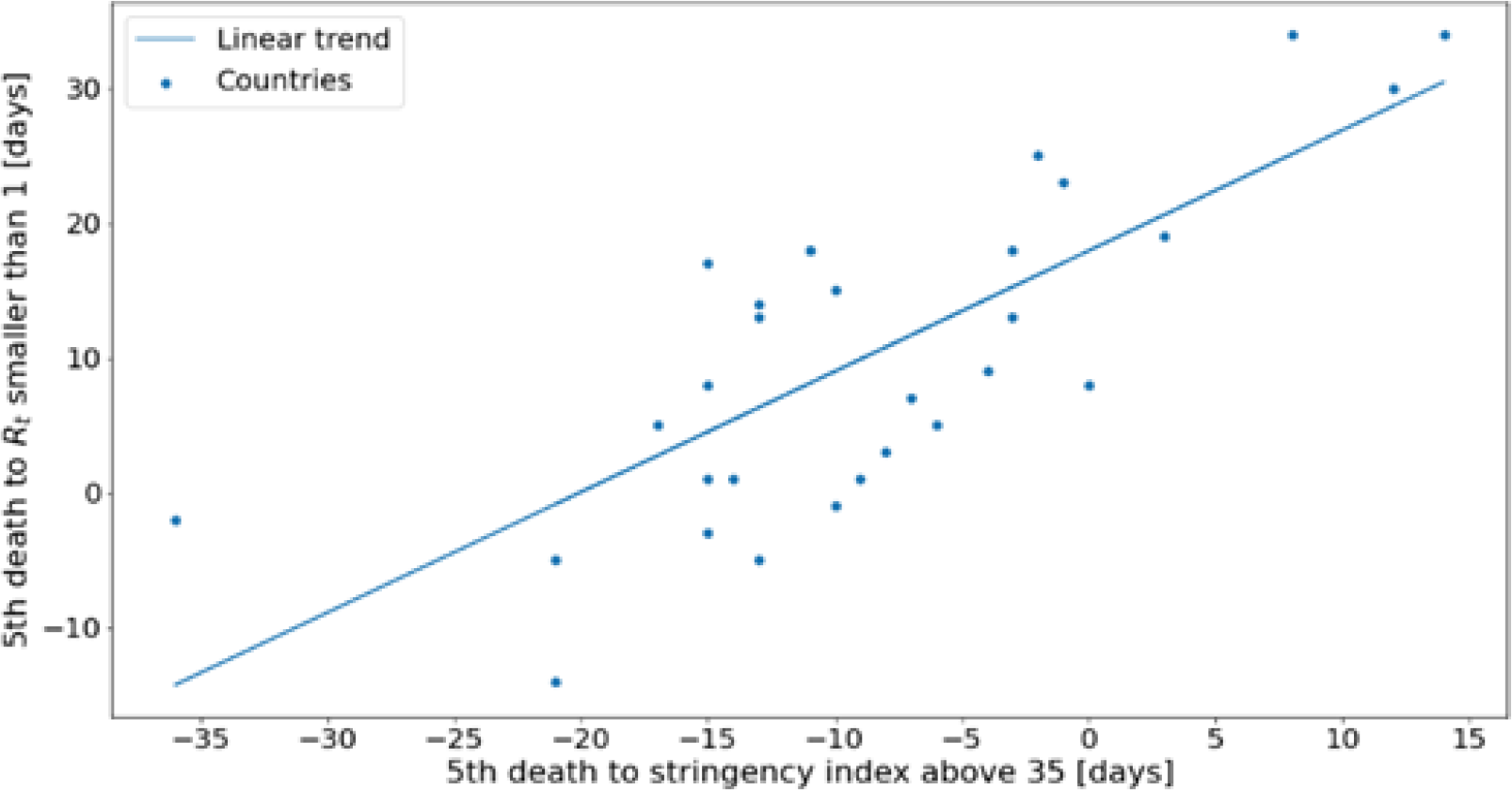
Period required to contain the epidemic as a function of the period to introduce initial restrictive measures.

The list of countries analyzed along dates characterizing the evolution of the epidemic and stringency index values are listed in Table 3 which is composed of three panels. The first panel includes the dates which were used to characterize the evolution of the pandemic in each country. The first columns of this panel is the date at which the 5^th^ death was observed, the 2^nd^ one when the stringency index reached a value of 35 and the third one includes the date at which the country managed to control the epidemic by reducing the reproduction number below one. The second panel shows the value of the stringency index when R_t_ was reduced below one. The last panel shows the period between the 5^th^ death and the stringency index reaching 35 or the reproduction number becoming smaller than one.

## 4 Discussion

Confirmed cases and death are widely available in the public domain, but to estimate the infection dates, the incubation period and the period between the onset of the symptoms and the person having a positive test or dying is required. The incubation period was initially derived on Chinese cases^22^ and it was assumed that this property is intrinsic to the virus and is therefore relevant for European countries. The period between the symptoms onset and a case being confirmed has been derived on Swiss patient^23^. The period between the symptoms onset and the death of the patient was derived on Chinese data^22^, but this period was not available for European patients. Based on 1430 Swiss cases, we found this period to have a mean of 15.3 days compared to 16.3 days in Linton et al.^22^. It was then assumed that this period was relevant for the European countries included in our study. It should be noted that this period is not only critical to understand how infections evolved at a given time, but is also very important to predict the occupancy of ICU units.

The method developed to estimate the effective reproductive number is based on the method developed by Cori et al.^13^. This method only requires the serial interval and an initial assumptions of the reproductive number. The advantage of the method presented in this report is that we are less reliant on the initial assumptions of the basic reproduction number. While Cori et al.^13^ assumes the prior is fixed in time, we constantly adapt it with new data. The initial assumptions will then converge to the computed reproductive number as more data are available. This is important as it is very difficult at the beginning of an epidemic to correctly evaluate the basic reproductive number R_0_ ^30^. In the future, our method will therefore be generalizable to new epidemics and provide reliable data at the start of the epidemic by being less reliant on the initial estimation of the basic reproduction number. However, as previous methods developed to estimate the reproduction number, our method is sensible to change in testing policy within a given country. Indeed these changes will most probably increase the number of positive cases without reflecting an increased seroprevalence within the society. It is also important to note that as there is a delay between the infection of an individual and the individual testing positive or dying, there is always a lag between the reproduction number measured today and the actual reproduction number.

Our analysis shows that when R_t_ reduced below one, the median severity of the measures for each category was important with a median stringency index of 83.6 out of 100. In addition the standard deviation of the index, which is equal to 13.5, shows that most countries required similar measures to control the epidemic. It is not possible to determine the impact of each individual measure as most countries took them in different order and often a given country took multiples ones at the same time, but the high stringency index reinforces the central idea that only important combinations of NPIs allow to control the pandemic. It is interesting to analyze the measure individually, not to determine their individual impact, but to determine which set of measures country had put in place when they successfully controlled the epidemic. If we look at the median restrictions when countries managed to control the epidemic, they were all at their maximum level apart from some exceptions on the closing of public transport as well as people being allowed to go out of with minimal daily exceptions. The two categories which had the strongest restrictions were the restrictions on public events and the school closing. All countries required cancelling public events apart from Sweden and Andorra which only recommended to cancel them. One limitation of the dataset used in this analysis is that it does not measure whether people have to wear mask either in public transport or in all closed environments. It would be important to include those data as more countries are introducing this type of measures to prevent the resurgence of the virus.

Our analysis also looked at the timing of the NPIs introduction. We found that there is a strong correlation (Pearson coefficient of 0.762) between the time at which NPIs were introduced and the time at which a country managed to control the epidemic with a reproductive number reducing below one. The United Kingdom can serve as an interesting example. The UK had initially planned to build “targeted herd immunity” delaying the introduction of restrictive measure. As a result of this delay, the UK were only able to contain the epidemic 30 days after the fifth death occurred in the country when the median time for the countries included in our analysis was of eight days. There are three outliers in our analysis being Andorra, Sweden and Iceland. Sweden has decided not to introduce a complete lockdown and stands with one of the highest daily death incidence in Europe (May 23^rd^ : Sweden - 5.34 deaths per million people per day; other European countries analyzed 0.82 on the same day ^25^).

A drawback of considering the evolution in the different countries at a national level and not at a regional one is that the heterogeneity of the spread of the virus is disregarded. To evaluate not only the effects of the NPIs but also the resurgence of localized clusters, it is important to look where the cases are located at a more local level. There is therefore a trade-off where the reproduction number is more reliable when evaluated on a larger amount of cases, but less representative as it does not take into account local disparities.

Our study on the impact of health measures focused on European countries but can be extended to other countries for which data on the daily incidence as well as the NPIs taken on a given day are available. To extend this study to a larger set of countries, it would however be necessary to adapt the period between the onset of the symptoms and a case being confirmed or the death of a patient. However, while a sensitivity analysis would be required to assert the influence of these parameters, the relatively small difference between the periods derived in Switzerland and in China (6.3%) in regards to the incertitude on the other parameters (daily incidence, serial interval) lets us believe that this factor is likely to play a marginal role if our analysis was to be extended to more countries.

Additional data could help refining our conclusions. First, hospitalizations data could be added as those would not be influenced by changes in testing policies within a given country. In addition, looking at the reproduction number within the different age groups could improve our understanding of the impacts of the different NPIs on these various groups. This information would be crucial to develop effective health policies protecting the most vulnerable while provoking minimal disruptions to the society and the economy.

## Data Availability

The incidence data for the confirmed and death cases were recovered from:Johns Hopkins University Center for Systems Science and Engineering, COVID-19 data reposi-tory, Center for Systems Science and Engineering (CSSE): https://github.com/CSSEGISandData/COVID-19 [accessed on 23/05/2020].
Data regarding the state interventions were retrieved from: Thomas Hale, Sam Webster, Anna Peth-erick, Toby Phillips, and Beatriz Kira. (2020). Oxford COVID-19 Government Response Tracker,Blavatnik School of Government: https://github.com/OxCGRT/covid-policy-tracker [accessed on 23/05/2020].
The daily incidence for the death cases per million people was retrieved from: Max Roser, HannahRitchie, Esteban Ortiz-Ospina and Joe Hasell, Coronavirus Pandemic (COVID-19), Our World inData, https://ourworldindata.org/coronavirus [accessed on 24/05/2020]
Data related to the period between a positive test and the death of an individual were retrieved from: Swiss Federal Office of Public Health (FOPH), Cas confirmes en laboratoire : distribution geographique, https://covid-19-schweiz.bagapps.ch/fr-1.html [accessed on 06/05/2020]

## 5 Conflict of Interest

The authors declare that the research was conducted in the absence of any commercial or financial relationships that could be construed as a potential conflict of interest.

## 6 Author Contributions

H.T. performed the analysis of the data as well as the redaction of the article

M.B. contributed to the analysis as well as the redaction of the article

A.R. reviewed the method used to analyze the data

C. G.-B. extracted the data used for the analysis

J.-P. G. reviewed the article

C.L. initiated the project and the research question and contributed to the redaction of the article

## 8 Data Availability Statement

The daily incidence for the death cases per million people was retrieved from: Max Roser, Hannah Ritchie, Esteban Ortiz-Ospina and Joe Hasell, Coronavirus Pandemic (COVID-19), Our World in Data, https://ourworldindata.org/coronavirus [accessed on 24/05/2020].

Data related to the period between a positive test and the death of an individual were retrieved from: Swiss Federal Office of Public Health (FOPH), Cas confirmés en laboratoire: distribution géographique, https://covid-19-schweiz.bagapps.ch/fr-1.html [accessed on 06/05/2020]

R_t_ was estimated using incidence data for confirmed cases and death published in the *COVID-19 Data Repository* by Johns Hopkins University (JHU CSSE) https://github.com/CSSEGISandData/COVID-19 [accessed on 23/05/2020].

Data regarding the various state interventions were retrieved from the *Coronavirus government response tracker* (OxCGRT) developed by the Blavatnik School of Government, Oxford University https://github.com/OxCGRT/covid-policy-tracker [accessed on 23/05/2020].

## Notes

### Competing Interest Statement

The authors have declared no competing interest.

### Funding Statement

No external funding was received.

### Summary of Updates

Corrected the order of the authors

